# Parametric identification and public health measures influence on the COVID-19 epidemic evolution in Brazil

**DOI:** 10.1101/2020.03.31.20049130

**Authors:** R.M. Cotta, C.P. Naveira-Cotta, P. Magal

## Abstract

A SIRU-type epidemic model is employed for the prediction of the COVID-19 epidemy evolution in Brazil, and analyse the influence of public health measures on simulating the control of this infectious disease. Since the reported cases are typically only a fraction of the total number of the symptomatic infectious individuals, the model accounts for both reported and unreported cases. Also, the model allows for a time variable functional form of both the transmission rate and the fraction of asymptomatic infectious that become reported symptomatic individuals, so as to reflect public health interventions, towards its control, along the course of the epidemic evolution. An analytical exponential behaviour for the accumulated reported cases evolution is assumed at the onset of the epidemy, for explicitly estimating initial conditions, while a Bayesian inference approach is adopted for parametric estimations employing the present direct problem model with the data from the known portion of the epidemics evolution, represented by the time series for the reported cases of infected individuals. The direct-inverse problem analysis is then employed with the actual data from China, with the initial phase of the data been employed for the parametric estimation and the remaining data being used for validation of the predictive capability of the proposed approach. The full dataset for China is then employed in another parameter identification, aimed at refining the values for the average times that asymptomatic infectious individuals and that symptomatic individuals remain infectious. Following this validation, the available data on reported cases in Brazil from February 15^th^ till March 29^th^, 2020, is used for estimating parameters and then predict the epidemy evolution from these initial conditions. As for the China analysis, the data for the reported cases in Brazil from March 30^th^ till April 23^rd^ are reserved for validation of the model. Finally, public health interventions are simulated, aimed at evaluating the effects on the disease spreading, by acting on both the transmission rate and the fraction of the total number of the symptomatic infectious individuals, considering time variable exponential behaviours for these two parameters, usually assumed constant in epidemic evolutions without intervention. It is demonstrated that a combination of actions to affect both parameters can have a more effective result in the control of the epidemy dynamics.

**NOMENCLATURE**

|  |  |
| --- | --- |
| $CR(t)$ | cumulative number of reported infectious symptomatic cases at time $t$ |
| $CU(t)$ | cumulative number of unreported infectious symptomatic cases at time $t$ |
| $DR(t)$ | daily number of reported infectious symptomatic cases at time $t$ |
| $f(t)$ | time-variable fraction of asymptomatic infectious who become reported symptomatic infectious |
| $f_0$ | initial fraction of asymptomatic infectious who become reported symptomatic infectious |
| $f_{max}$ | second value of fraction of asymptomatic infectious who become reported symptomatic infectious |
| $f_{max2}$ | third value of fraction of asymptomatic infectious who become reported symptomatic infectious |
| $I(t)$ | number of asymptomatic infectious individuals at time $t$ |
| $I_0$ | asymptomatic infectious individuals at initial time $t_0$ |
| $N$ | time in days of application of the first public health intervention that changes transmission rate |
| $N_2$ | time in days of application of the second public health intervention that changes transmission rate |
| $N_f$ | time in days for application of the first public health intervention that changes the fraction of asymptomatic infectious that become reported symptomatic infectious |
| $N_{f2}$ | time in days for application of the second public health intervention that changes the fraction of asymptomatic infectious that become reported symptomatic infectious |
| $NP$ | total number of parameters in estimation algorithm |
| $M$ | total number of measured data used in the estimation |
| $R(t)$ | number of reported symptomatic infectious individuals at time $t$ |
| $R_0$ | reported symptomatic infectious individuals at initial time $t_0$ |
| $\mathbf{P}$ | vector of parameters under estimation ($P_1, P_2, \dots, P_{NP}$) |
| $S(t)$ | number of individuals susceptible to infection at time $t$ |
| $S_0$ | number of individuals susceptible to infection at initial time $t_0$ |
| $t$ | time variable, in days |
| $t_0$ | beginning date of the epidemic, in days |
| $U(t)$ | number of unreported symptomatic infectious individuals at time $t$ |
| $U_0$ | unreported symptomatic infectious individuals at initial time $t_0$ |
| $\mathbf{Y}$ | vector of measured data ($Y_m$ at time $t_m, m = 1, \dots, M$) |

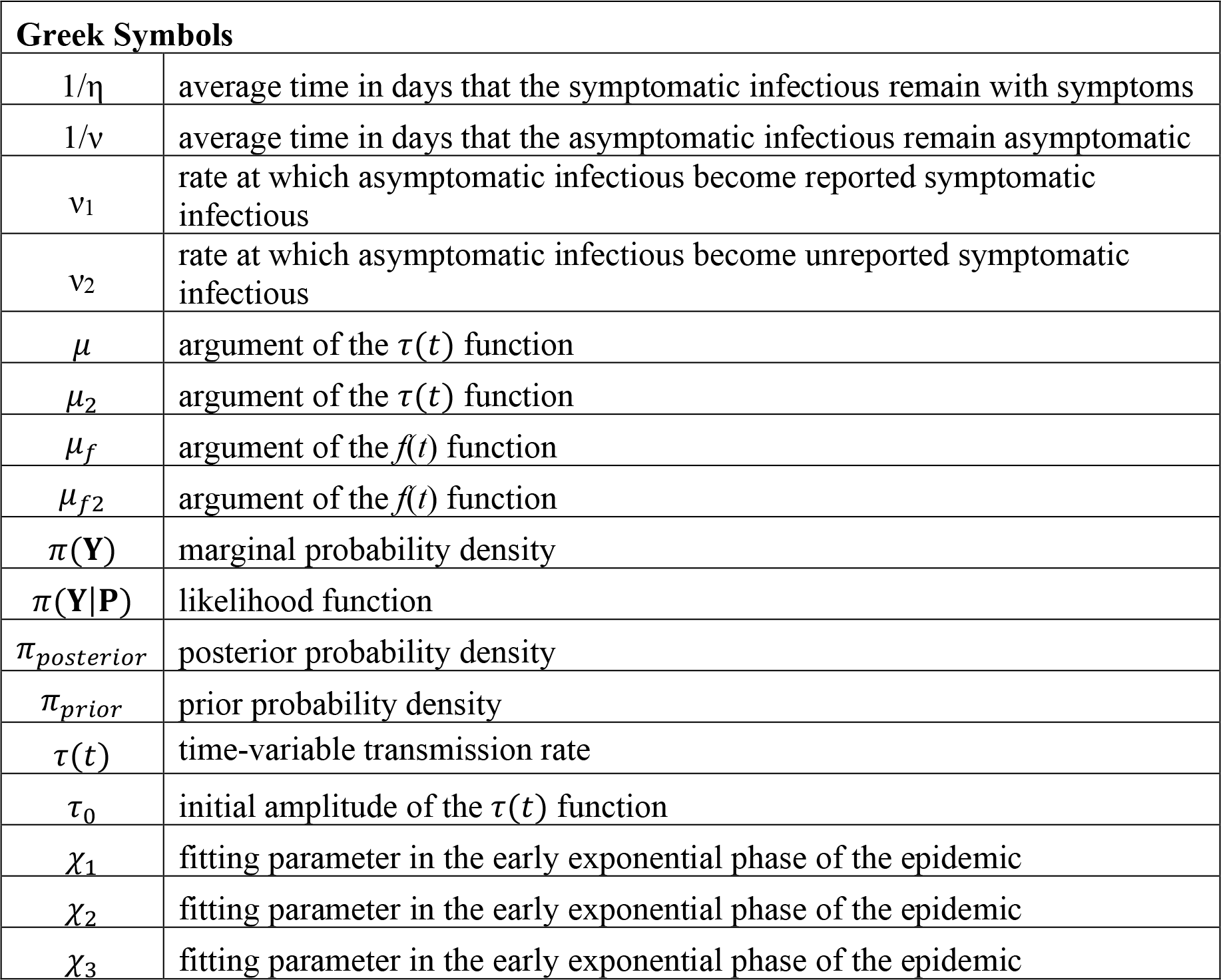

## INTRODUCTION

A new human coronavirus started spreading in Wuhan, China, by the end of 2019, and turned into a pandemic disease called COVID-19 as declared by the World Health Organization on March 11^th^, 2020. The affected countries and cities around the world have been reacting in different ways, towards locally controlling the disease evolution. These measures include general isolation through quarantine and massive testing for focused isolation, with varying degrees of success so far, as can be analysed from the limited data available. Naturally, China offers the longest time series on reported infected cases and the resulting effects of combining different public health interventions. As of March 26^th^, 2020, there were no reports in China of further internal contaminations, and all the new cases are associated with infected individuals that (re)entered in the country. Despite the apparent success of the interventions in China, each region or country might require a specific combination of measures, due to demographic spatial distribution and age structure, health system capabilities, and social-economical characteristics. In this sense, it urges to have a mathematical model that would allow for the simulation of such possible interventions on the epidemic evolution within the following weeks or months. This article presents a collaborative research effort towards the construction of an epidemic evolution prediction tool, which combines direct and inverse problem analysis and is both reliable and easy to implement and execute, initially motivated by offering some insight into the control of COVID-19 within Brazil.

The classical susceptible-infectious-recovered (SIR) model describes the transmission of diseases between susceptible and infective individuals and provides the basic framework for almost all epidemic models. At the onset of the coronavirus epidemy in China, there were some initial studies for the prediction of its evolution and the analysis of the impact of public health measures [1], which however did not consider in the modelling the presence of unreported infectious individuals cases, which are in practice inherent to this process. The present work is first based on the SIRU-type model proposed in [2], which deals with the epidemic outbreak in Wuhan by introducing the unreported cases in the modelling, and evaluating the consequences of public health interventions. It was a direct application of previous developments [3,4] on the fundamental problem of parameter identification in mathematical epidemic models, accounting for unreported cases. This same modelling approach was more recently employed in the analysis of the epidemic outbreak in different countries, including China, South Korea, Germany, Italy, and France [5-7]. Besides identifying unreported cases, this simple and robust model also allows for introducing a latency period and a time variable transmission rate, which can simulate a public health orientation change such as in a general isolation measure. In addition, an analytical exponential behaviour is assumed for the accumulated reported cases evolution along an initial phase just following the onset of the epidemy, which, upon fitting of the available data, allows for the explicit analytical estimation of the transmission rate and the associated initial conditions required by the model.

Here, the SIRU-type model in [2-7] is implemented for the direct problem formulation of the COVID-19 epidemic evolution, adding a time variable parametrization for the fraction of asymptomatic infectious that become reported symptomatic individuals, a very important parameter in the public health measure associated with massive testing and consequent focused isolation. The same analytical identification procedure is maintained for the required initial conditions, as obtained from the early stage exponential behaviour. However, a Bayesian inference approach is here adopted for parametric estimation, employing the Markov Chain Monte Carlo method with the Metropolis-Hastings sampling algorithm [8-12]. At first, the goal of the inverse problem analysis was estimating the parameters associated with the transmission rate and the fraction of asymptomatic infectious that become reported symptomatic individuals, which can be quite different in the various regions and countries and may also vary according to the public health measures. Then, in light of the success in this parametric identification, an extended estimation was also employed which incorporates the average time the asymptomatic infectious are asymptomatic and the average time the infectious stay in the symptomatic condition, due to the relative uncertainty on these parameters in the literature. The proposed approach was then applied to the data from China, first by taking just the first portion of these data points in the estimation, while using the second portion to validate the model using the estimated parameters with just the first phase of the epidemy evolution, and second by employing the whole time series in the MCMC estimation procedure, thus identifying parameters for the whole evolution period. This second estimation was particularly aimed at refining the data for the average times that asymptomatic infectious individuals and that symptomatic individuals remain infectious. Upon validation of the approach through the data for China, we have proceeded to the analysis of the epidemic dynamics in Brazil, employing about 36 days (February 15^th^ till March 29^th^) of collected information on reported infected individuals. First, the available data was employed in the parametric estimation, followed by the prediction of the epidemy evolution in Brazil. For this purpose, the following 20 days (from March 31^st^ to April 19^th^) were reserved to be used in the validation of the proposed model for the COVID-19 evolution in Brazil. Finally, we have explored the time variation of both the transmission rate and the fraction of asymptomatic infectious that become reported symptomatic individuals, so as to reflect public health interventions, in simulating possible government measures, as described in what follows.

## DIRECT PROBLEM

The implemented SIRU-type model [2-7] is given by the following initial value problem:

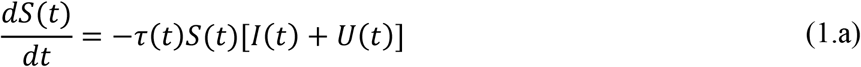

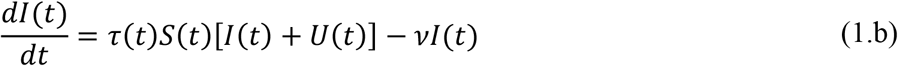

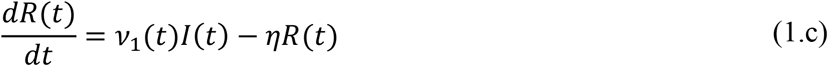

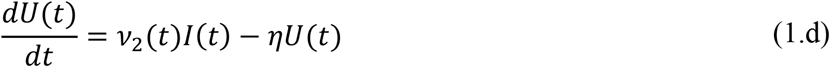

where,

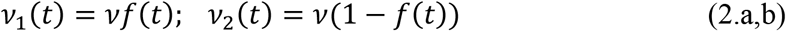

with initial conditions

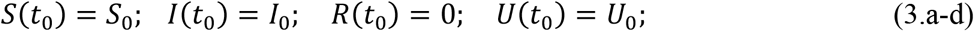

Here, t_0_ is the beginning date of the epidemic in days, S(t) is the number of individuals susceptible to infection at time t, I(t) is the number of asymptomatic infectious individuals at time t, R(t) is the number of reported symptomatic infectious individuals (i.e., symptomatic infectious with severe symptoms) at time t, and U(t) is the number of unreported symptomatic infectious individuals (i.e., symptomatic infectious with mild symptoms) at time t. Asymptomatic infectious individuals I(t) are infectious for an average period of 1/ν days. Reported symptomatic individuals R(t) are infectious for an average period of 1/η days, as are unreported symptomatic individuals U(t). We assume that reported symptomatic infectious individuals R(t) are reported and isolated immediately, and cause no further infections. The asymptomatic individuals I(t) can also be viewed as having a low-level symptomatic state. All infections are acquired from either I(t) or U(t) individuals. The fraction f(t) of asymptomatic infectious become reported symptomatic infectious, and the fraction 1-f(t) become unreported symptomatic infectious. The rate asymptomatic infectious become reported symptomatic is ν_1_(t) = f(t)v, the rate asymptomatic infectious become unreported symptomatic is ν_2_(t) = (1 -f(t)) ν, where ν_1_(t) + ν_2_(t) = ν. The transmission rate, *τ*(t), is also allowed to be a time variable function along the evolution process. Figure 1 below illustrates the infection process as a flow chart.

**Figure 1.**
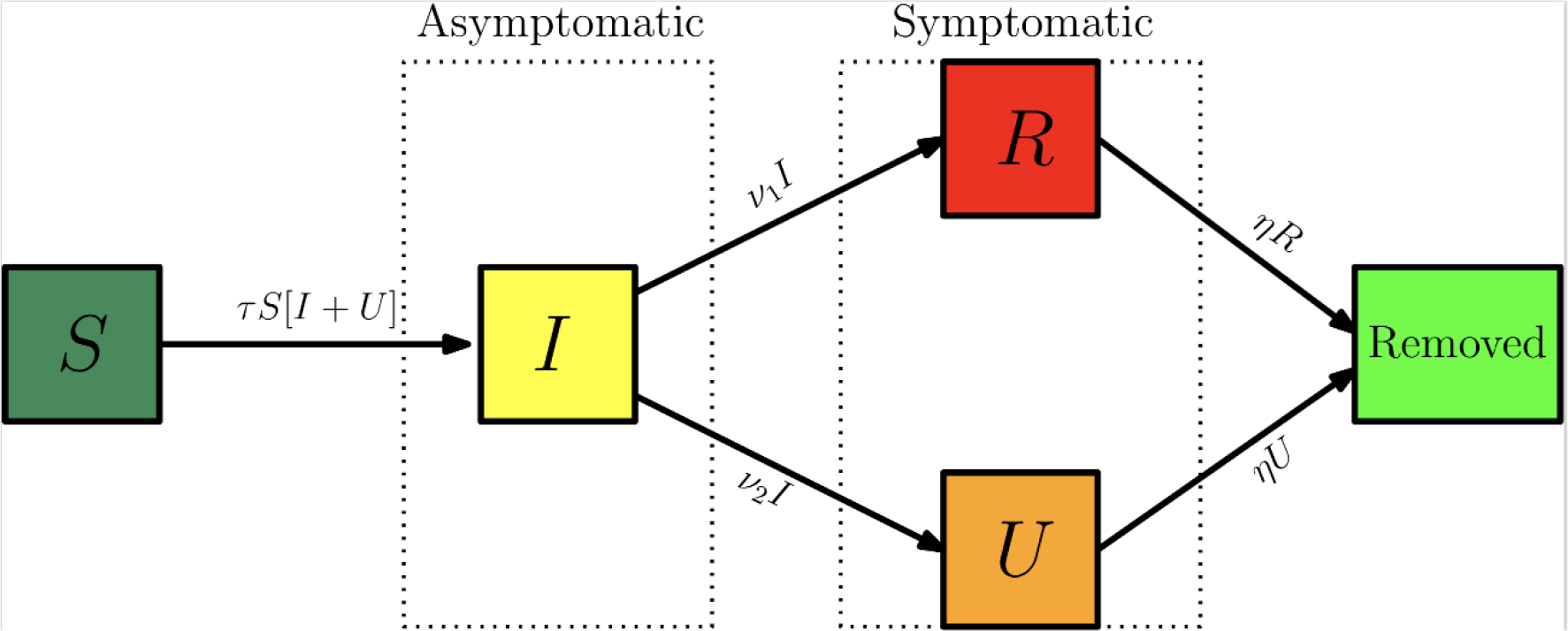
– Flow chart illustrating the infection path process [3].

The time variable coefficients, *τ*(t) and f(t), are chosen to be expressed as:

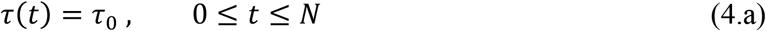

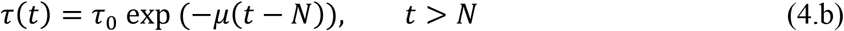

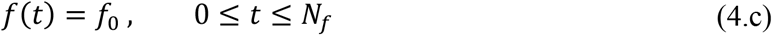

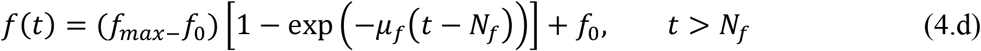

These parametrized functions are particularly useful in interpreting the effects of public health interventions. For instance, the transmission rate, *τ*(t), is particularly affected by a reduced circulation achieved through a general isolation or quarantine measure, while the fraction f(t) of asymptomatic infectious that become reported, thus isolated, cases can be drastically increased by a massive testing measure with focused isolation. In the above relations, *μ* is the attenuation factor for the transmission rate, N is the time in days for application of the public health intervention to change transmission rate, *μ_f_* is the argument of the *f*(*t*) variation between the limits (*f*_0_,*f_max_*). The first time variable function has been previously considered, while the second one has been introduced in the present work, so as to allow for the examination of combined measures.

The cumulative number of reported cases at time t, *CR*(*t*), which is the quantity offered by the actual available data, and the a priori unknown cumulative number of unreported cases, *CU*(*t*), are given by:

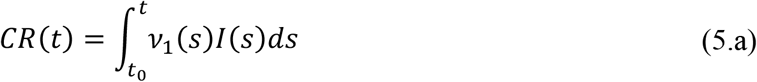

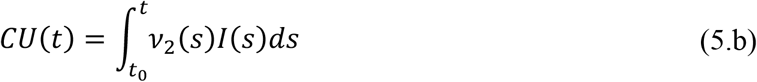

The daily number of reported cases from the model, *DR*(*t*), can be obtained by computing the solution of the following equation:

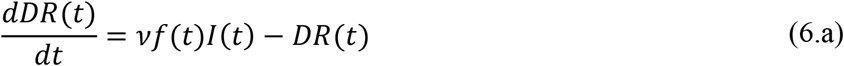

with initial conditions

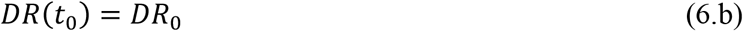

## INVERSE PROBLEM

Inverse problem analysis is nowadays a common practice in various science and engineering contexts, in which the groups involved with experimental data and numerical simulation collaborate so as to obtain the maximum information from the available data, towards the best possible use of the modelling for the problem under study. Here, as mentioned in the introduction, we first review an analytical parametric identification described in more details in [4-7], that from the initial phases of the epidemic evolution allows to explicitly obtain the unknown initial conditions of the model, while offering a reliable estimate for the transmission rate at the onset of the epidemy. Nevertheless, even after these estimates, a few other parameters in the model remain uncertain, either due to the specific characteristics of the physical conditions or response to the epidemy in each specific region, or due to lack of epidemiological information on the disease itself. Therefore, an inverse problem analysis was undertaken aimed at estimating the main parameters involved in the model, as summarized in Table 1 below. First, for the dataset on the accumulated reported cases for China, the focus is on the parametrized time variation of the transmission rate (*τ*_0_ and *μ*) and the fraction of asymptomatic infectious that become reported (*f*_0_), in this case assumed constant, followed by an effort to refine the information on the average times (1/ν and 1/η) through a simultaneous estimation of the five parameters. Then, employing the dataset for Brazil just up to March 29^th^, the parametrized time variation of the transmission rate (*τ*_0_ and *μ*) and the fraction of asymptomatic infectious that become reported *f*(*t*), assumed time-variable, are estimated by parametrization of an abrupt variation that requires just the estimation of *f_max_* and *N_f_*.

**Table 1.** – Summary of the estimated parameters on each inverse problem analysis.

| China |  |  |
| --- | --- | --- |
| Case | Parameter under estimation | Data range used in the Inv. Prob. |
| case 1: CH3p | $f_0, \mu, \tau_0$ | January 19<sup>th</sup> up to February 17<sup>th</sup> |
| case 2: CH5p-full | $f_0, \mu, \tau_0, 1/\nu, 1/\eta$ | January 19<sup>th</sup> up to April 16<sup>th</sup> |
| Brazil |  |  |
| case 3: BR5p | $f_0, \mu, \tau_0, f_{max}, N_f$ | February 25<sup>th</sup> up to March 29<sup>th</sup> |

The statistical inversion approach here implemented falls within the Bayesian statistical framework [8-12], in which (probability distribution) models for the measurements and the unknowns are constructed separately and explicitly, as shall be briefly reviewed in what follows.

As explained in previous works employing this model [4-7], it is assumed that in the early phase of the epidemic, the cumulative number of reported cases grows approximately exponentially, according to the following functional form:

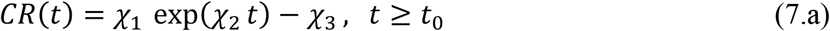

After fitting this function to the early stages of the epidemic evolution, one may extract the information on the unknown initial conditions, in the form [4-7]:

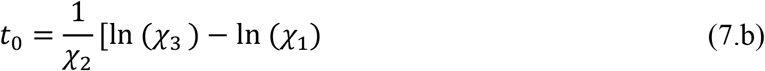

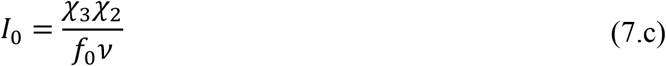

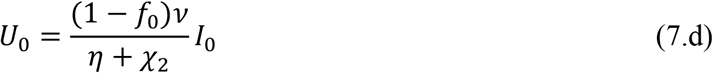

In addition, an excellent estimate for the initial transmission rate can be obtained from the same fitted function, in the form:

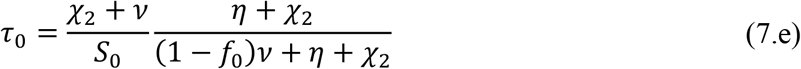

Also, the basic reproductive number for this initial phase model is estimated as:

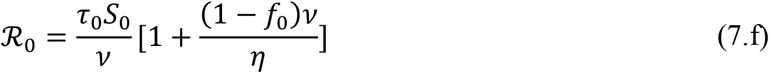

The statistical approach for the solution of inverse problems here adopted employs the Metropolis-Hastings algorithm for the implementation of the Markov chain Monte Carlo (MCMC) method [8-9]. The MCMC method is used in conjunction with the numerical solution of the ordinary differential system, eqs.(1-3), for estimating the remaining model parameters. Consider the vector of parameters appearing in the physical model formulation as:

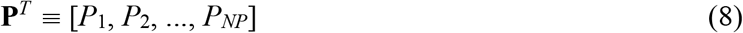

where *NP* is the number of parameters. For estimating **P**, we assume that a vector of measured data is available (**Y**) containing the measurements *Y_m_* at time *t_m_*, *m* = 1,…, *M*. Bayes’ theorem can then be stated as [8-9]:

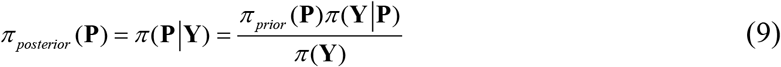

where *π_posterior_*(**P**) is the posterior probability density, that is, the conditional probability of the parameters **P** given the measurements **Y**, *π*_prior_(**P**) is the prior density, that is, the coded information about the parameters prior to the measurements, *π* (**Y**|**P**) is the likelihood function, which expresses the likelihood of different measurement outcomes **Y** with **P** given, and *π*(**Y**) is the marginal probability density of the measurements, which plays the role of a normalizing constant. If different *prior* probability densities are assumed for the parameters, the posterior probability distribution may not allow an analytical treatment. In this case, Markov chain Monte Carlo (MCMC) methods are used to draw samples of all possible parameters, and thus inference on the posterior probability becomes inference on the samples [8-9]. The main merit of the MCMC method is about providing a picture of the posterior distribution, without solving the mathematical integrals in Bayes’ rule. The idea is to approximate the posterior distribution by a large collection of samples of values. This method is especially suitable when it is unfeasible to yield an analytical solvable posterior distribution and/or a large space of parameters is involved, allowing one to do Bayesian inference even in rich and complex models. The idea behind the Metropolis-Hasting sampling algorithm is illustrated below, and these steps should be repeat until it is judged that a sufficiently representative sample has been generated.

1. Start the chain with an initial value, that usually comes from any prior information that you may have;
2. Randomly generate a proposed jump aiming that the chain will move around and efficiently explores the region of the parameter space. The proposal distribution can take on many different forms, in this work a Gaussian random walk was employed, implying that the proposed jumps will usually be near the current one;
3. Compute the probability of moving from the current value to the proposed one. Candidates moving to regions of higher probability will be definitely accepted. Candidates in regions of lower probability can be accepted only probabilistically. If the proposed jump is rejected, the current value is tally again. For more details on theoretical aspects of the Metropolis-Hastings algorithm and MCMC methods and its application, the reader should refer to [8-12].

## RESULTS AND DISCUSSION

### Model Validation: China

Before proceeding to the analysis of the COVID-19 epidemic evolution within Brazil, which is the major concern in the present contribution, the need was felt in validating the proposed direct-inverse problem analysis approach. In this sense, due to the availability of the largest dataset on this pandemic, we have chosen to use the information from China in terms of the accumulated confirmed infectious cases. The data for China was extracted from [6], complemented by the most recent data from [13] from January 1^st^ up to April 17^th^, 2020. The exponential fit for the early phase of the China CR(t) dataset provided the estimates of the three parameters, *χ*_1_ = 0.14936, *χ*_2_ = 0.37680, *χ*_3_ = 1.0, from which we have estimated *t*_0_ = 5.046. The remaining data for the initial conditions, *I*_0_ and *U*_0_, and the early stage transmission rate, *τ*_0_, are in fact recalculated from within the MCMC algorithm, since the changing values of *f* will affect them, according to eqs. (7.c-e). The average times in the model were taken as 1/ν=7 and 1/η=7 days and the isolation measures were taken at N=25 days [6]. First, experimental data from China from the period of January 19^th^ up to February 17^th^ was employed in demonstrating the estimation of three parameters, *f*_0_,*μ*,and *τ*_0_, assuming there is no significant time variation in the function *f*(*t*) (*μ_f_* = 0). In the absence of more informative priors, uniform distributions were employed for all three parameters under estimation. Table 2 presents the prior information and the initial guesses for the parameters. If the initial guesses were used to predict the CR(t) behavior, an over-estimation of the accumulated reported infected individuals would occur, especially in the long term, as can be noticed in Figure 1, confirming the need for a proper parameter estimation.

The central tendency (mean value) of the posteriors here sampled, after neglecting the first 20,000 burning in states of the chain, are called the estimated values. Both the estimated values and their 99% confidence intervals are presented in Table 3. It should be mentioned that these values are fairly close to those employed in [6], where τ_0_ was estimated as 4.51×10^-8^. Once a value of *f_0_* = 0.8 was assumed, which means that 20% of symptomatic infectious cases go unreported, it led to a good agreement with the data by taking *μ*=0.139 in [6].

**Table 2.** – Prior distributions and initial guesses for the parameters to be estimated *f*_0_, *μ*, and *τ*_0_ (Wuhan, China).

| Case 1: CH3p |  |  |  |
| --- | --- | --- | --- |
| Param. | Prior distribution | Initial Guess |  |
| $f$ | $U[0, 1]$ | 0.5 | estimated |
| $\mu$ | $U[0, 5]$ | 0.1 | estimated |
| $\tau_0$ | $U[0, 1 \times 10^{-6}]$ | $4.478 \times 10^{-8}$ | estimated |
| $S_0$ | $11.0 \times 10^6$ | | fixed |
| $t_0$ | 5.04617 | | fixed |
| $1/\nu$ | 7 days | | fixed |
| $1/\eta$ | 7 days | | fixed |
| $N$ | 25 | | fixed |
| $\chi_1$ | 0.14936 | | fixed |
| $\chi_2$ | 0.37680 | | fixed |
| $\chi_3$ | 1.0 | | fixed |

**Figure 1.**
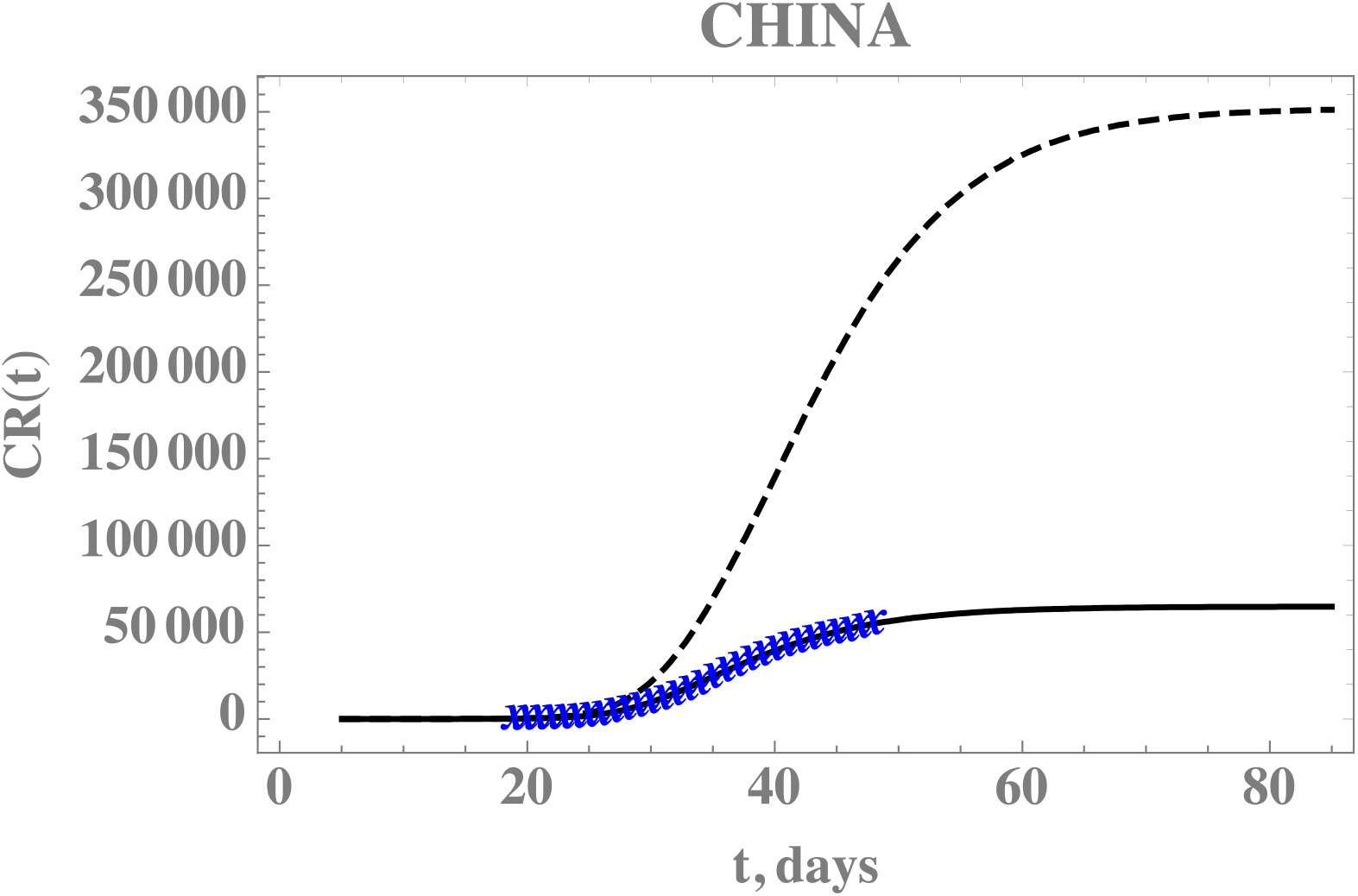
– Comparison of the theoretical model for CR(t) with the initial guesses from Table 2 (black dashed line), against the model prediction with the estimated values from Table 3 (solid black line), and actual data from China from January 19^th^ to February 17^th^ (blue cross) -- Case 1: CH3p

Figure 1 also demonstrates the adherence of the model with the data within this portion of the dataset, once the estimated values in Table 3 are employed in the direct problem solution, as can be seen from the excellent agreement between the estimated CR(t) (solid line) and the experimental data from China (blue stars). The desired model validation is illustrated in Figure 2, confirming the excellent agreement of China’s full dataset (period from January 19th till April 16th) with the mathematical model predictions, after adopting the estimated values for the parameters in Table 3. It should be recalled that non-informative priors were adopted for the three parameters, as presented in Table 2, and except for the transmission rate, when eq.(7.e) provides an excellent initial guess, the remaining guesses were completely arbitrary.

**Figure 2.**
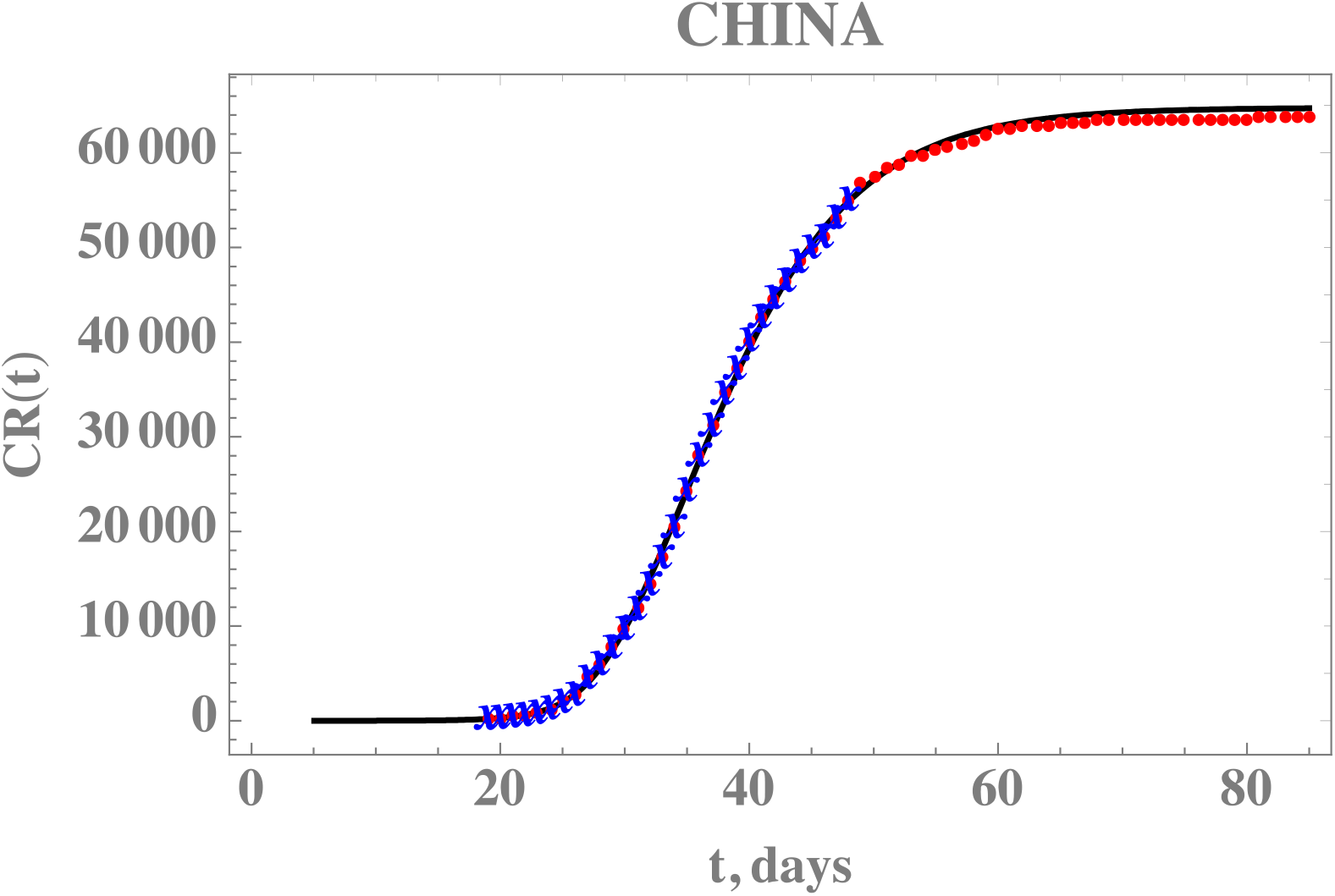
– Comparison of the theoretical model for CR(t) with the three estimated parameter values from Table 3 (solid line), against the dataset for China from January19^th^ to February17^th^ (blue stars) and from February18^th^ to April 16^th^ (red dots) -- Case 1: CH3p

**Table 3.** – Estimated values and 99% confidence intervals for three parameters, *f*_0_, *μ*, and *τ*_0_ (Wuhan, China).

| Case 1: CH3p |  |  |
| --- | --- | --- |
| Parameter | Estimated values | 99% confidence interval |
| $f$ | 0.780719 | [0.77956, 0.7818] |
| $\mu$ | 0.135635 | [0.135219, 1.136153] |
| $\tau_0$ | $4.47793 \times 10^{-8}$ | $[4.47793 \times 10^{-8}, 4.47793 \times 10^{-8}]$ |

Although the present estimated parameters have led to a good prediction of the second phase of the China epidemic evolution data, there are still uncertainties associated with the average times here assumed both equal to 7 days, according to [6]. This choice was based on early observations of the infected asymptomatic and symptomatic patients in Wuhan, but more recent studies have been refining the information on the epidemic evolution and the disease itself, such as in [14-17]. For this reason, we have also implemented a statistical inverse analysis with the full dataset of China, but now seeking the estimation of five parameters, so as to simultaneously estimate the average times (1/ν and 1/η). Both uniform and Gaussian distributions were adopted for the two new parameters, with initial guesses of 1/ν=7 days and 1/η=7 days, and N=25 days, as employed in [6]. Table 4 presents the prior information and the initial guesses for the parameters.

**Table 4.**
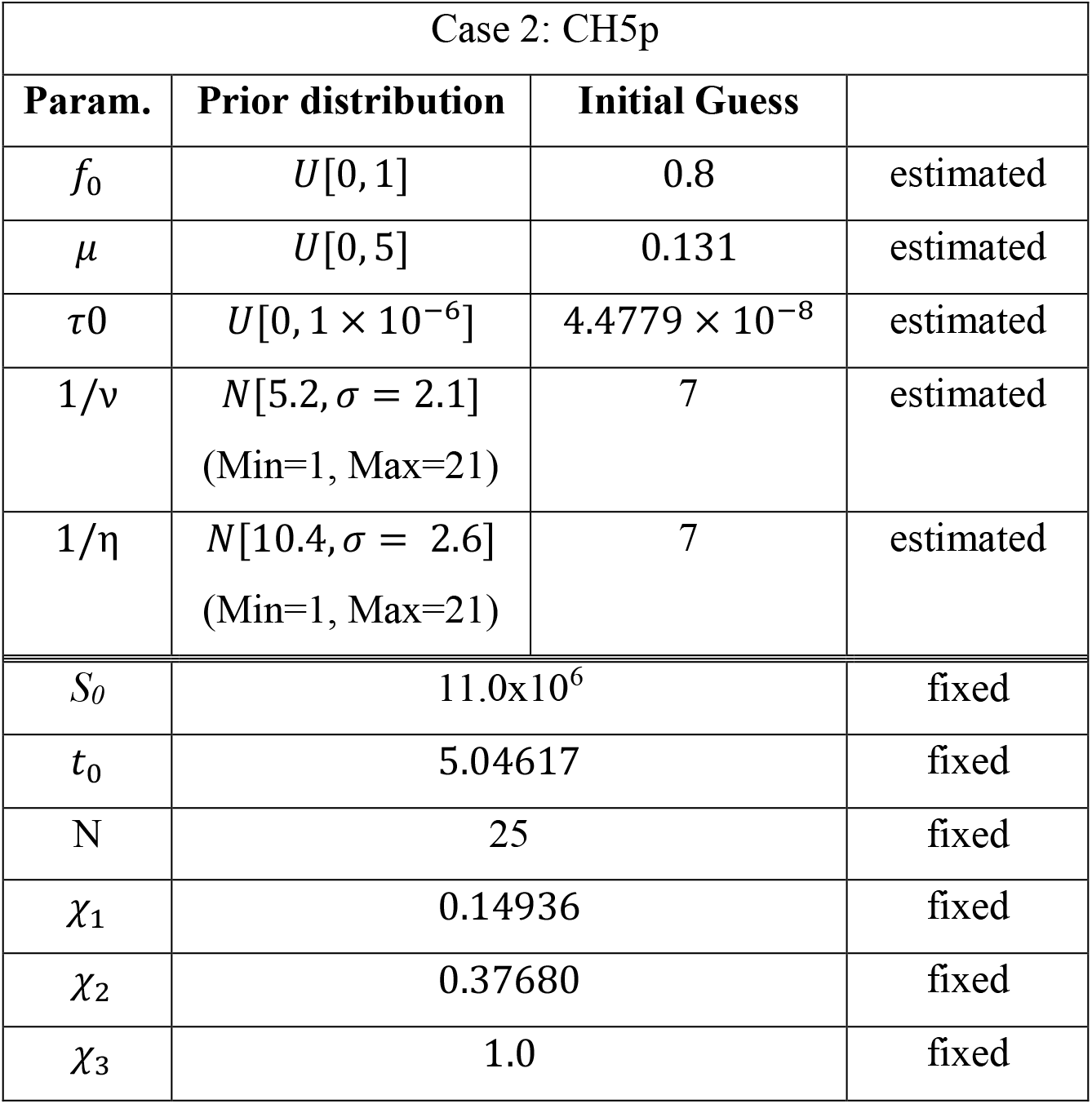
– Prior distributions and initial guesses for the 5 parameters to be estimated, *f*_0_, *μ*, *τ*_0_,1/ν and 1/η, (Wuhan, China)

| Case 2: CH5p |  |  |  |
| --- | --- | --- | --- |
| Param. | Prior distribution | Initial Guess |  |
| $f_0$ | $U[0, 1]$ | 0.8 | estimated |
| $\mu$ | $U[0, 5]$ | 0.131 | estimated |
| $\tau_0$ | $U[0, 1 \times 10^{-6}]$ | $4.4779 \times 10^{-8}$ | estimated |
| $1/\nu$ | $N[5.2, \sigma = 2.1]$<br>(Min=1, Max=21) | 7 | estimated |
| $1/\eta$ | $N[10.4, \sigma = 2.6]$<br>(Min=1, Max=21) | 7 | estimated |
| $S_0$ | 11.0x10<sup>6</sup> | | fixed |
| $t_0$ | 5.04617 | | fixed |
| N | 25 |  | fixed |
| $\chi_1$ | 0.14936 | | fixed |
| $\chi_2$ | 0.37680 | | fixed |
| $\chi_3$ | 1.0 | | fixed |

Table 5 provides the estimated values and 99% confidence intervals for all five parameters, with Gaussian priors for the two average times with data obtained from [14,17], after neglecting the first 15,000 burn in states of the chain. The most affected parameter in comparison with the previous estimates is the average time 1/η, which is also the one with widest confidence interval. This behaviour is also evident from the Markov chains for this parameter, now simultaneously estimated. Figure 3 compares the theoretical predictions with the model incorporating the five estimated parameters as in Table 5, against the full CR(t) dataset for China, confirming the improved agreement. The 99% confidence interval bounds for this predicted behavior is lso shown in Fig.3, bounded by the gray lines.

**Table 5.**
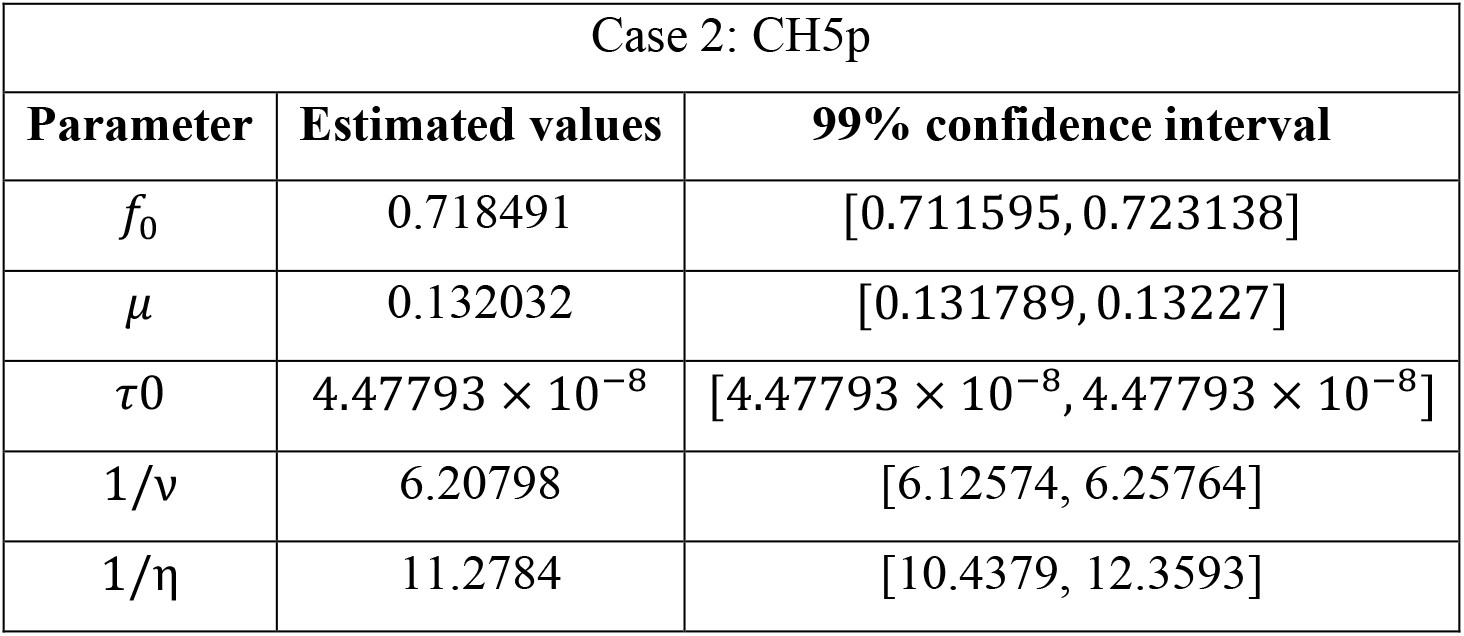
– Estimated values and 99% confidence intervals for five parameters, *f*_0_, *μ*, *τ*_0_,1/ν and 1/η (Wuhan, China).

**Figure 3.**
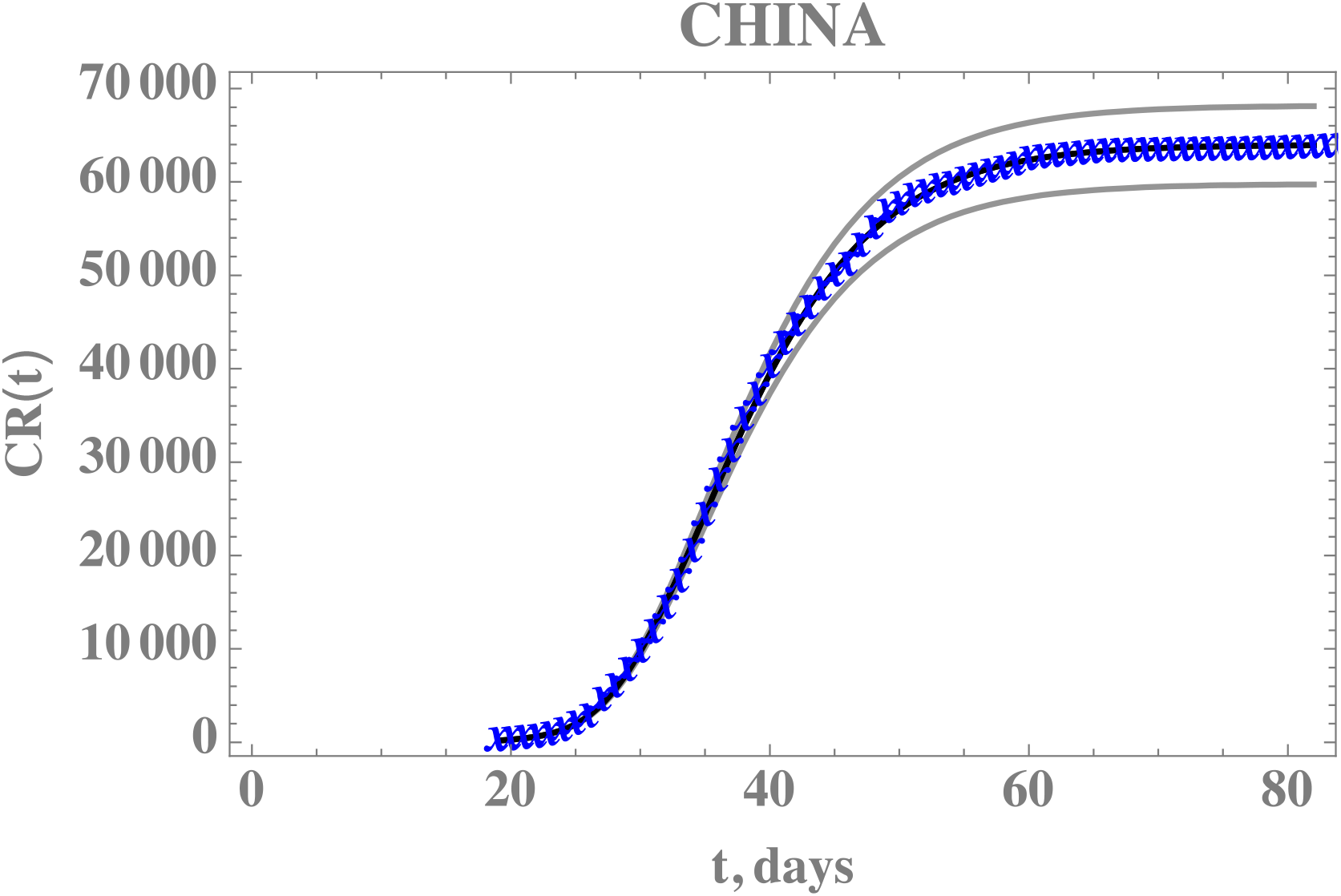
– Comparison of the theoretical model for CR(t) with the five estimated parameter values (black solid line) and 99% confidence intervals (gray lines), against the complete dataset for China from January 19th up to April 16th (blue cross) – Case 2: CH5p

### Model Application: Brazil

The CR(t) data for the accumulated reported infectious in Brazil, from February 25^th^, when the first infected individual was reported, up to April 23rd, is presented in the Appendix. Similarly to the previous example with the data from China, a portion of the available data on accumulated reported infectious was employed in the model parameters estimation, up to March 29^th^. Then, the second portion of the data, from March 30^th^ up to April 23rd, was utilized in validating the constructed model.

First, the exponential phase of the evolution was fitted, taking the data from day 10 to 25, yielding the estimates of the three parameters, *χ*_1_ = 0.42552, *χ*_2_ = 0.293696, *χ*_3_ = 3.2335, from which we have estimated *t*_0_ = 6.9051. The remaining data for the initial conditions, *I*_0_ and *U*_0_, and the early stage transmission rate, *τ*_0_, are in fact recalculated from within the MCMC algorithm, since the changing values of *f*_0_ will affect them, according to eqs. (7.c-e). The average times in the model were taken as 1/ν= 6.20798 days and 1/η= 11.2784 days, which were obtained from the MCMC simulation on the full dataset for China (Table 5), as discussed in the previous section.

The Brazilian government took isolation measures starting on N=21 days, which was enforced throughout the country. Also, there were initially only 30,000 exam kits available, and an additional 30,000 were later acquired, but till mid April at least, the resulting rather small ration of testing per million inhabitants in Brazil and the retardation in the exam results confirmation due to a centralized operation, has caused a perceptible change in the data structure for the reported infectious cases, which can only be represented by a time varying function *f*(*t*). The progressive reduction on the number of executed exams of the symptomatic individuals and the delay of the results availability, has certainly affected the partition of reported to unreported cases by the end of this period covered by the present dataset. Therefore, the more general model including the time variation of the partition *f*(*t*), eqs.(4.c,d), is here implemented for a more refined inverse problem analysis. It is then expected that a reduction on the f value can be identified (*f_max<_f*_0_), with an abrupt variation on the exponential behaviour, here assumed as a sharp functional time dependence (large *μ_f_*). Therefore, a statistical inverse problem analysis is undertaken, this time for estimating five parameters, namely, *f*_0_, *μ*, *τ*_0_, *f_max_*, and *N_f_* (Case 3 – BR5p) aimed at enhancing the overall agreement with the CR(t) data behaviour, with a likely reduction on the partition of the reported and unreported infectious cases.

With uniform distributions for all five parameters, guided by the previous estimates for the first three parameters, and arbitrary guesses for *f_max_*, and *N_f_*, the prior distributions and initial guesses for the 5 parameters are presented in Table 6 and the five estimated quantities, after neglecting the first 80,000 burning in states of the chain are shown in Table 7, together with the 99% confidence interval for each parameter

**Table 6.** – Prior distribution and initial guesses for the 5 parameters to be estimated, *f*_0_, *μ*, *f_max_*, *N_f_* (Brazil)

| Case 3 - BR5p |  |  |  |
| --- | --- | --- | --- |
| Param. | Prior distribution | Initial Guess |  |
| $f_0$ | $U[0, 1]$ | 0.300 | estimated |
| $\mu$ | $U[0, 5]$ | 0.04 | estimated |
| $\tau_0$ | $U[0, 1 \times 10^{-6}]$ | $1.66755 \times 10^{-9}$ | estimated |
| $f_{max}$ | $U[0, 1]$ | 0.165 | estimated |
| $N_f$ | $U[10, 35]$ | 30.5 | estimated |
| $1/\nu$ | 6.20798 days | | fixed |
| $1/\eta$ | 11.2784 days | | fixed |
| $S_0$ | $211.3 \times 10^6$ | | fixed |
| $t_0$ | 6.90514 | | fixed |
| $N$ | 21 | | fixed |
| $\mu_f$ | 10 | | fixed |
| $\chi_1$ | 0.42552 | | fixed |
| $\chi_2$ | 0.293696 | | fixed |
| $\chi_3$ | 3.2335 | | fixed |

Figure 4 presents the predicted evolution of the accumulated reported infectious cases in Brazil, CR(t), from February 25^th^ up to April 23rd, plotted as the black dashed line. Also shown in this figure are the red dots in the first portion of the available data, up to March 29^th^, which were employed in the estimation of the parameters in Table 7 that compose the present model. In addition, the blue dots represent the second portion of the available data from March 30^th^ till April 23rd, that were not employed in the parametric estimation, but saved for the present validation. It is clear that the built model has an excellent predictive feature, reproducing the epidemic evolution up to the available date at the time of this work submission, with a mean relative error of 5.8% during this phase.

**Figure 4.**
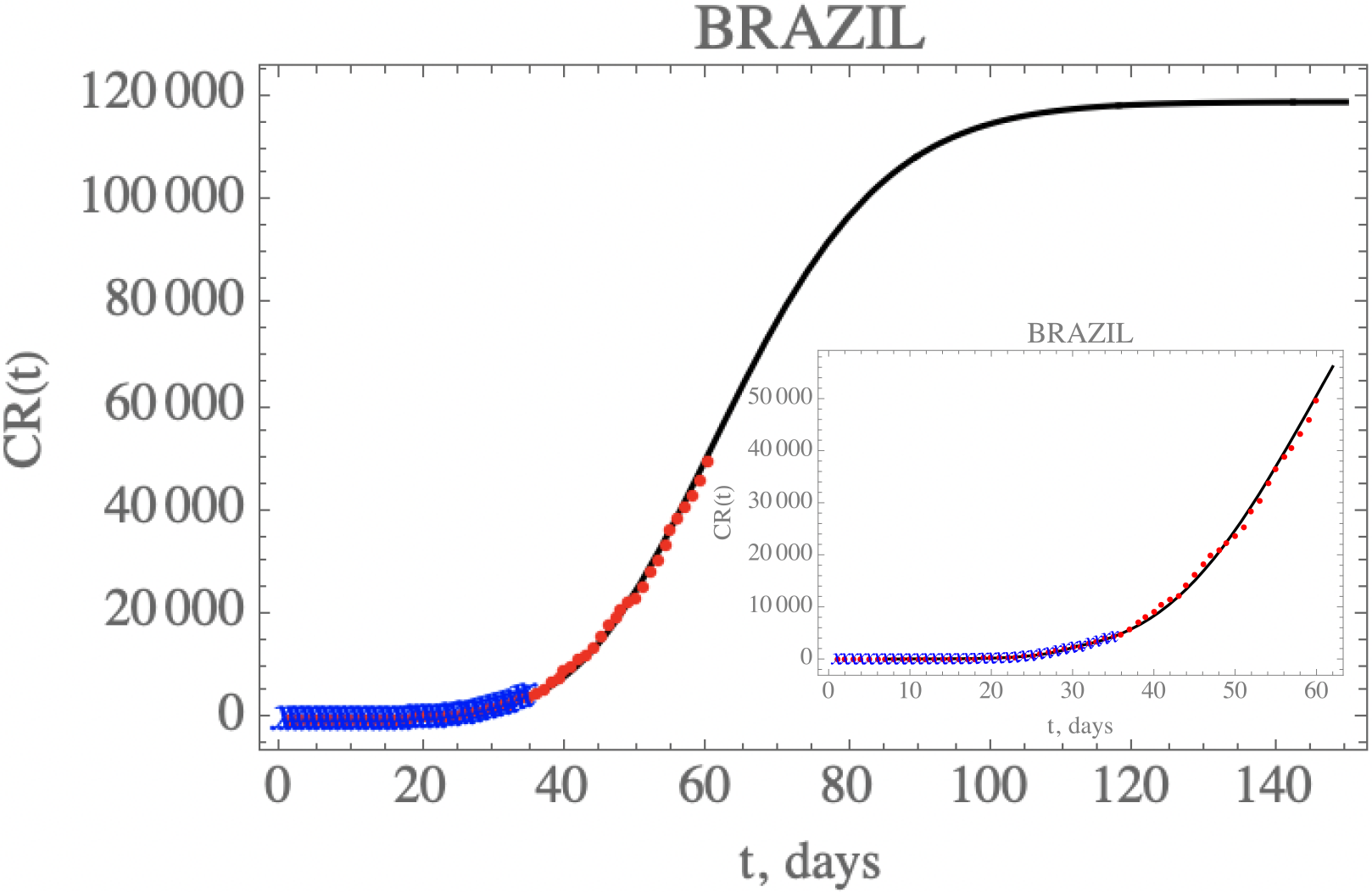
– Prediction of the accumulated reported infectious, CR(t), with the five estimated parameter values from the available dataset for Brazil from February 25^th^ up to March 29^th^ (blue cross) and validated with the data up to April 23^rd^ (red dots).

**Table 7.** – Estimated values and 99% confidence intervals for five parameters, *f*_0_, *μ*, *f_max_*, and *N_f_* (Brazil).

| Case 3 – BR5p |  |  |
| --- | --- | --- |
| Parameter | Estimated values | 99% confidence interval |
| $f_0$ | 0.303671 | [0.302624, 0.304697] |
| $\mu$ | 0.0389639 | [0.0388438, 0.0390961] |
| $\tau_0$ | $1.66755 \times 10^{-9}$ | $[1.66755 \times 10^{-9}, 1.66755 \times 10^{-9}]$ |
| $f_{max}$ | 0.156734 | [0.156146, 0.157217] |
| $N_f$ | 30.4197 | [30.3522, 30.4915] |

One can see the marked reduction on the *f*(*t*) parameter from the estimates in Table 7, which results in the increase of the unreported to reported infectious cases, as is shown in Figure 5 for CR(t) and CU(t) predictions up to 150 days. Clearly, the reduction on the testing, and thus on the isolation of reported infectious individuals, leads to an impressive increase on the total number of infected symptomatic individuals after 150 days (752,888 cases), including unreported (633,698) and reported cases (119,190). Both the reported and unreported infectious individuals curves, R(t) and U(t), show a peak at around the 70^th^ day (May 3^rd^).

**Figure 5.**
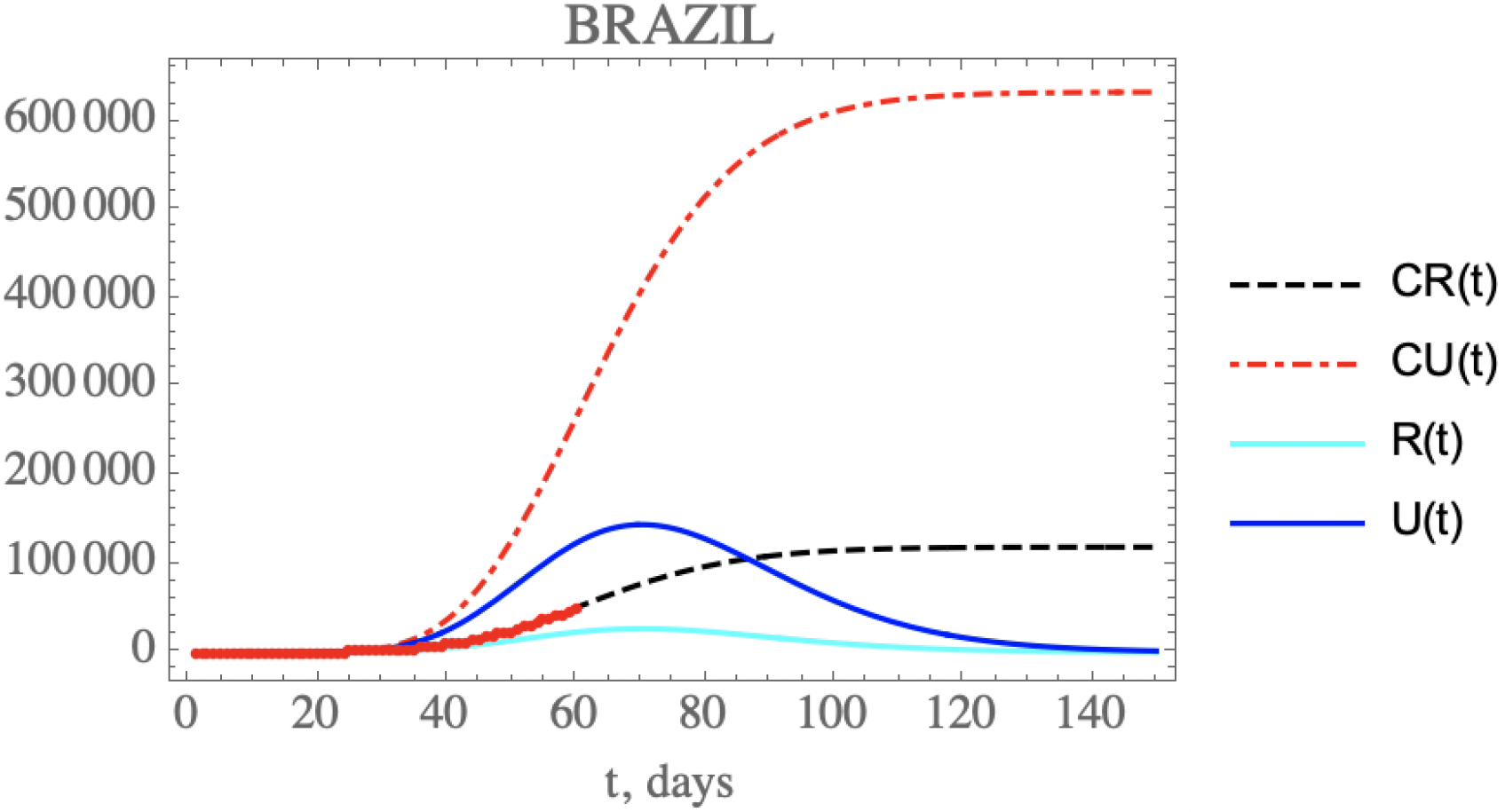
– Comparison of the theoretical model for CR(t) (black dashed curve), CU(t) (red dashed curve), R(t) (cyan solid curve) and U(t) (blue solid curve) with the five estimated parameter values from the available dataset for Brazil from February 25^th^ up to March 29^th^. (red dots show 60^th^ available data of CR(t) up to April 23^rd^)

### Scenarios analyses: Brazil

Next, the model constructed with this parametric estimation is employed in the prediction of the COVID-19 evolution in Brazil under different hypothesis. Five scenarios were here explored: (i) the present public health interventions remain active, with the same transmission rate decay; (ii) a stricter social distancing and/or prevention measures are implemented from now on, further reducing the transmission rate decay; (iii) an attenuation on the social distancing and/or prevention measures, leading to a more mild reduction on transmission rate; (iv) an increment on the fraction of reported cases, through a more intensive blood testing, for instance, leading to more unreported cases to become reported ones, thus isolating them earlier; (v) a combination of public health measures acting on reducing the transmission rate, though with some relaxation of the social distancing, but simultaneously increasing the conversion factor of unreported to reported cases.

Table 8 summarizes both the fixed and variable parameter values adopted for these five scenarios. The additional public health interventions simulated in the above scenarios act on either the time variation of the transmission coefficient or on the reported to unreported partition coefficient. Since April 23rdcorresponds to t=59 days, the parametric changes are assumed to start at a chosen date further ahead, in the present case t=64 days (*N*_2_ = *N_f_*_2_ = 64), and the scenarios analysis are undertaken by acting on either or both coefficients, *τ*(*t*) and *f*(*t*), according to the following parametrizations:

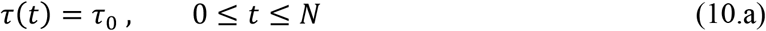

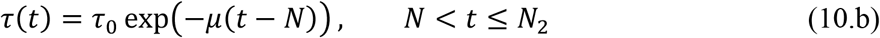

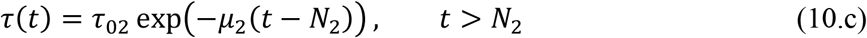

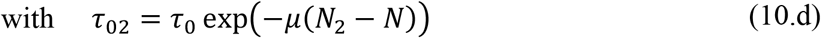

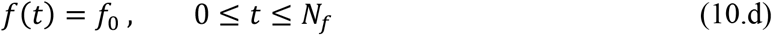

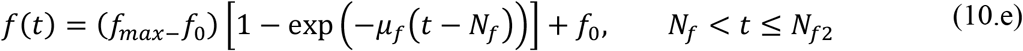

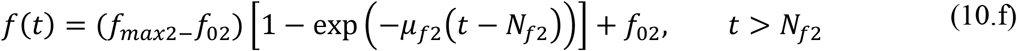

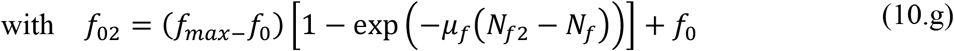

In the first scenario, it is assumed that no additional public health interventions are implemented, other than those already reflected by the data up to April 23rd, which would then be fully maintained throughout the control period, and the epidemics should evolve from the present stage, under the parameters above identified. Figure 5 has already shown the evolution of the accumulated and instantaneous reported, CR(t) and R(t), unreported, CU(t) and U(t), infectious individuals, up to 150 days. Due to the fairly low value of f(t) starting with *f*_0_ ≈ 0.30 and reaching *f_max_* ≈ 0.16, the accumulated number of unreported infectious cases is quite high, as already discussed. No predictions on casualties are here proposed, since these are highly dependent on age structure, social-economical conditions, and health system response.

**Table 8.** – Input data in each scenario for epidemic evolution in Brazil

| Fixed parameters |  |  |  |  |  |
| --- | --- | --- | --- | --- | --- |
| $f_0$ | 0.303671 | | | | |
| $\mu$ | 0.0389639 | | | | |
| $N$ | 21 | | | | |
| $\tau_0$ | $1.66755 \times 10^{-9}$ | | | | |
| $f_{max}$ | 0.156734 | | | | |
| $\mu_f$ | 10 | | | | |
| $N_f$ | 30.4197 | | | | |
| $1/\nu$ | 6.20798 days | | | | |
| $1/\eta$ | 11.2784 days | | | | |
| $S_0$ | $211.3 \times 10^6$ | | | | |
| $t_0$ | 6.90514 | | | | |
| $\chi_1$ | 0.42552 | | | | |
| $\chi_2$ | 0.293696 | | | | |
| $\chi_3$ | 3.2335 | | | | |
| Changing parameters: (64<sup>th</sup> – 150<sup>th</sup> day) |  |  |  |  |  |
| Scenario | (i) | (ii) | (iii) | (iv) | (v) |
| $\mu_2$ | 0 | 0.0779278 | 0.019482 | 0 | 0.019482 |
| $N_2$ | - | 64 | 64 | - | 64 |
| $f_{max2}$ | - | - | - | 0.607342 | 0.607342 |
| $\mu_{f2}$ | 0 | 0 | 0 | 10 | 10 |
| $N_{f2}$ | - | - | - | 64 | 64 |

Next, the second scenario explores the implementation of more strict distancing and sanitary habits to further reduce the transmission rate by assuming, after day N_2_=64 (eq.10.c), by doubling the value of *μ* here identified, thus around, *μ*_2_=0.0779, still well below that achieved in China (0.132). The time variable transmission rate is then computed from eq.(10.c) after t > N_2_. The changes in the accumulated reported and unreported cases, as shown in Figure 6, are quite significant. The predicted number of unreported symptomatic infectious cases is now much lower reaching after 150 days around 545,324 individuals, while the reported cases should reach 102,764 individuals, with an impressive reduction to a total of around 648,088 infectious symptomatic cases. The predicted evolution of the reported infectious cases would then show a peak at around t=69 days.

**Figure 6.**
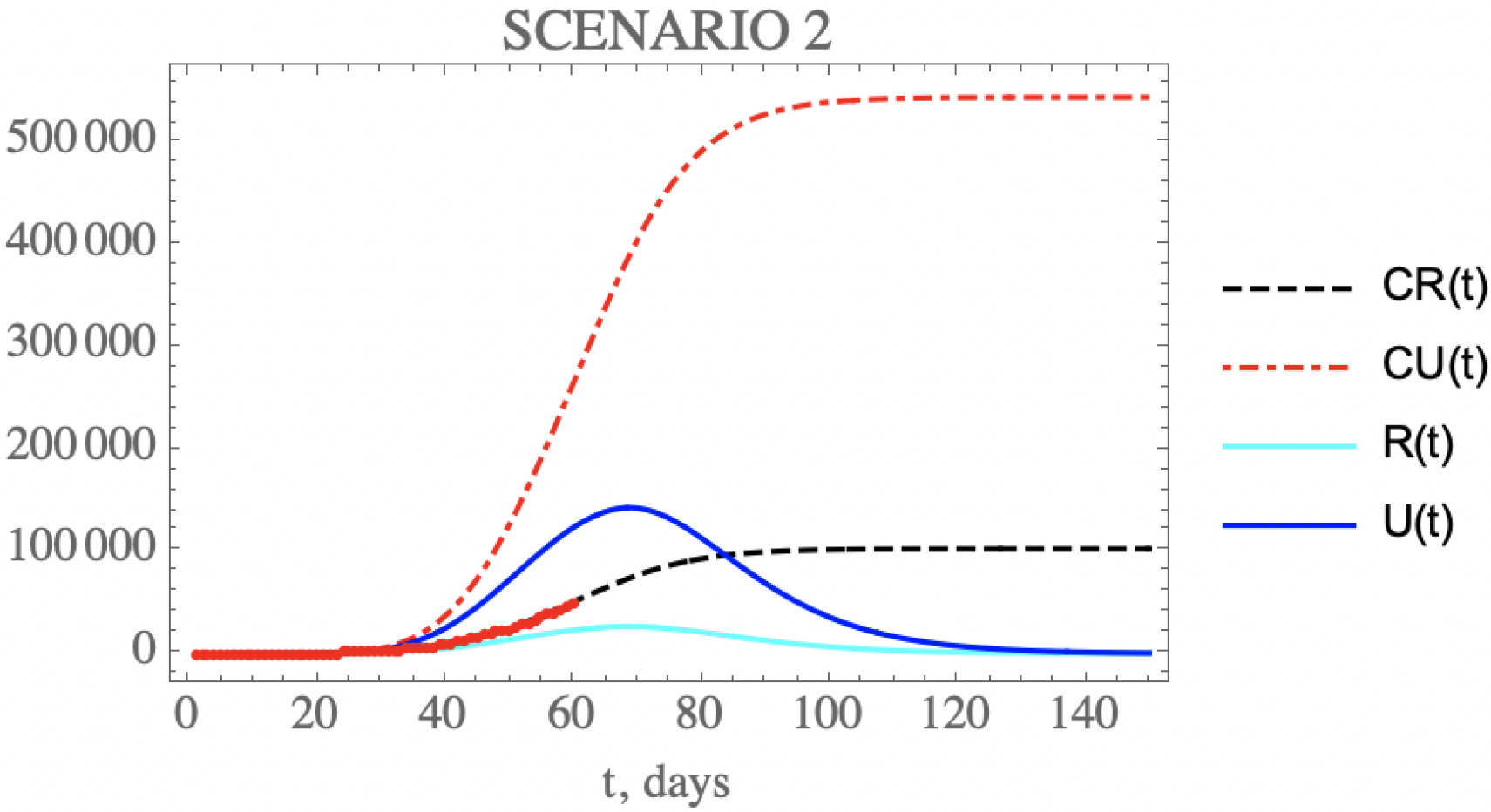
– Scenario (ii) predictions for CR(t) (black dashed curve), CU(t) (red dashed curve), R(t) (cyan solid curve) and U(t) (blue solid curve) with the five estimated parameter values from the available dataset for Brazil from February 25^th^ up to March 29^th^. (red dots show available data of CR(t) up to April 23^rd^).

Through the third scenario, one can predict the consequences of mildly relaxing the public health measures that affect transmission rate, for instance through reduction of isolation measures. This is simulated here by reducing the identified transmission rate attenuation factor, by assuming, after day N_2_=64, half the value of *μ* here identified, thus around, *μ*_2_=0.0195. The changes in the accumulated and instantaneous reported and unreported symptomatic cases, as shown in Figure 7, are worse than in the base scenario (i), Figure 5. The predicted number of accumulated unreported infectious cases is now higher reaching after 150 days around 765,612 individuals, while the reported cases would reach 143,708 individuals, with an increase to a total of around 909,319 infectious symptomatic cases. The predicted evolution of the instantaneous reported infectious cases would then show a peak at around t=72 days.

**Figure 7.**
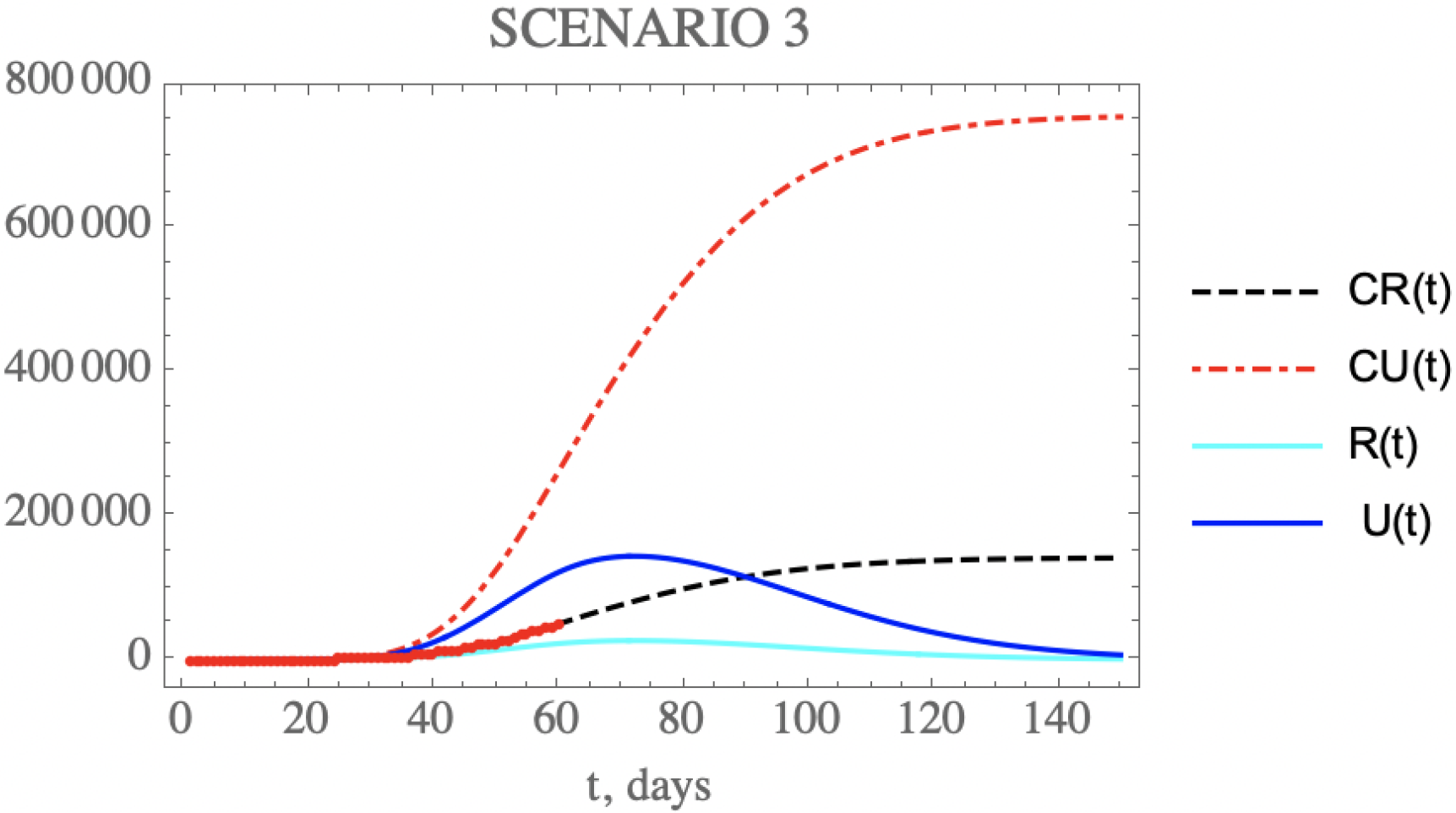
– Scenario (iii) predictions for CR(t) (black dashed curve), CU(t) (red dashed curve), R(t) (cyan solid curve) and U(t) (blue solid curve) with the five estimated parameter values from the available dataset for Brazil from February 25^th^ up to March 29^th^. (red dots show available data of CR(t) up to April 23rd).

Besides acting on the transmission rate along time, public health measures may also be effective in reducing the ratio of reported to unreported infectious case, since the reported cases are, according to the model, directly isolated and thus interrupting the contamination path, as analyzed in the fourth scenario. For instance, one may double the initial fraction of reported and unreported infectious cases parameter, *f*_0_, to reach *f_max_*_2_ = 0.607, after *N_f_*_2_ = 64 *days*, somehow closer to the value previously obtained from the China dataset. Therefore, Figure 8 shows the behavior of CR(t), R(t) and CU(t), U(t), which according to the value of *μ_f_*_2_ = 10, occurring after the day *N_f_*_2_ = 64, leads to the crossing of instantaneous reported and unreported cases, R(t) and U(t), that can be observed. The predicted number of unreported infectious cases would now reach, after 150 days, around 441,949 individuals, while the reported cases should reach 241,931 individuals, with an also marked reduction to a total of around 683,880 infectious cases. The predicted evolution of the reported infectious cases would then show a peak at around t=76 days. Although this peak value is higher than for the base case scenario (i), before further public health intervention, a number of these are of mild symptomatic cases that were moved from the unreported to the reported cases evolution, thus isolated earlier.

**Figure 8.**
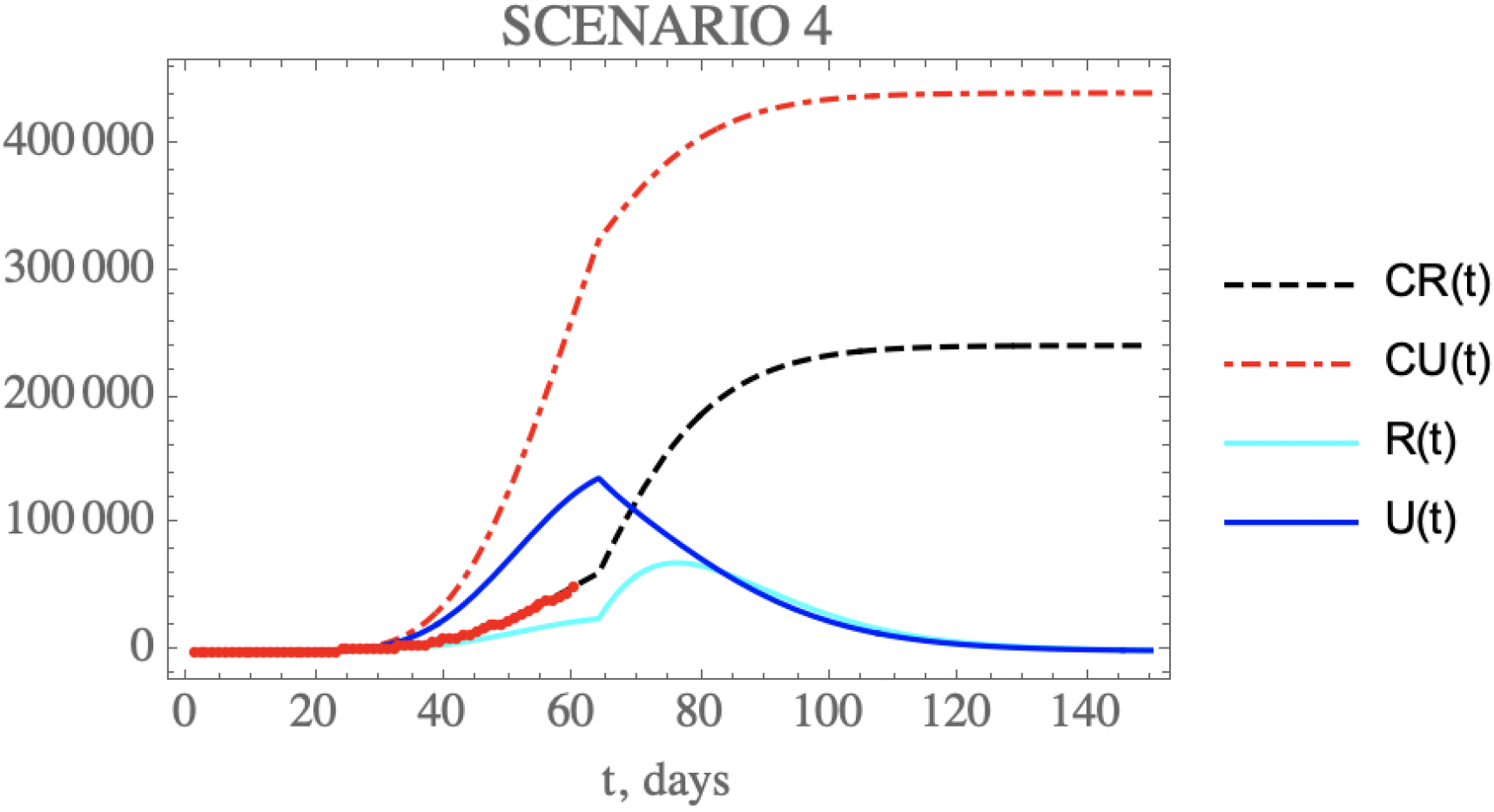
– Scenario (iv) predictions for CR(t) (black dashed curve), CU(t) (red dashed curve), R(t) (cyan solid curve) and U(t) (blue solid curve) with the five estimated parameter values from the available dataset for Brazil from February 25^th^ up to March 29^th^. (red dots show available data of CR(t) up to April 23^rd^).

In the fifth scenario, the combination of public health measures affecting both the transmission rate and the conversion factor of unreported to reported cases is analyzed for Brazil. Let us consider after day N_2_=64, some relaxation of social distancing leading to half of the *μ* value here identified, thus around, *μ*_2_=0.0195, and simultaneously doubling the fraction of reported and unreported infectious cases, to become *f_max_*_2_ = 0.607, also after *N_f_*_2_ *=* 64 *days*, with *μ_f_*_2_=10. The changes in the accumulated and instantaneous reported and unreported cases are shown in Figure 9. The predicted number of unreported infectious cases is now reaching after 150 days around 471,320 individuals, while the reported cases should reach 287,360 individuals, with a total of 758,680 infectious symptomatic cases, less than 1% increase with respect to the base case. The predicted evolution of the daily reported infectious cases would then show a peak at around t=78 days. Again, though this peak value is higher than for the base case, before the public health improvements, a number of these are of mild symptomatic cases that were moved from the unreported to the reported cases evolution, thus moved to monitored isolation earlier, and not necessarily requiring hospitalization. In overall terms, the results for scenario (v) are not markedly different from those for the base scenario (i), thus offering a perspective of combining the social distancing relaxation measures with more intensive testing to reach a similar final effect.

**Figure 9.**
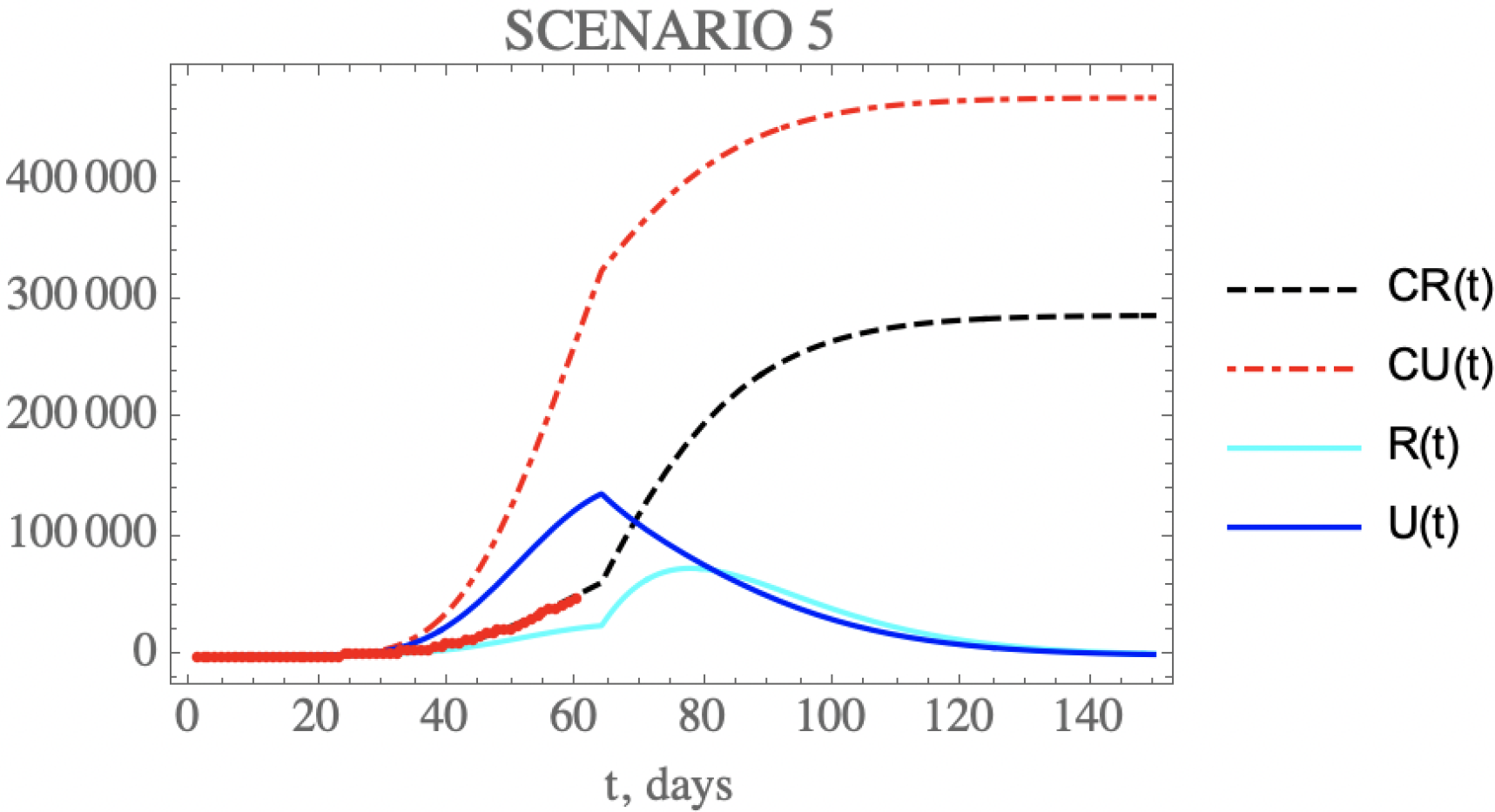
– Scenario (v) predictions for CR(t) (black dashed curve), CU(t) (red dashed curve), R(t) (cyan solid curve) and U(t) (blue solid curve) with the five estimated parameter values from the available dataset for Brazil from February 25^th^ up to March 29^th^. (red dots show available data of CR(t) up to April 23^rd^).

Figures 10.a,b combine the data on accumulated reported and unreported infectious symptomatic individuals, respectively, for the predictions provided through the five scenarios here considered. Clearly, scenario (ii), which involves further restrictions on social distancing and sanitary habits, and scenario (iv) which involves more intensive testing while maintaining the present public health actions, lead to the smaller accumulated values of symptomatic individuals in the long term, while the plain relaxation of social distancing, without any other intervention, would result in the largest number of infected symptomatic cases, either reported or unreported. On the other hand, when examining the curves for scenarios 1 and 5, it is clear that the proper combination of public health interventions, which would involve relaxation of social distancing and intensification of testing, could result in similar results as a more strict quarantine process.

**Figures 10.**
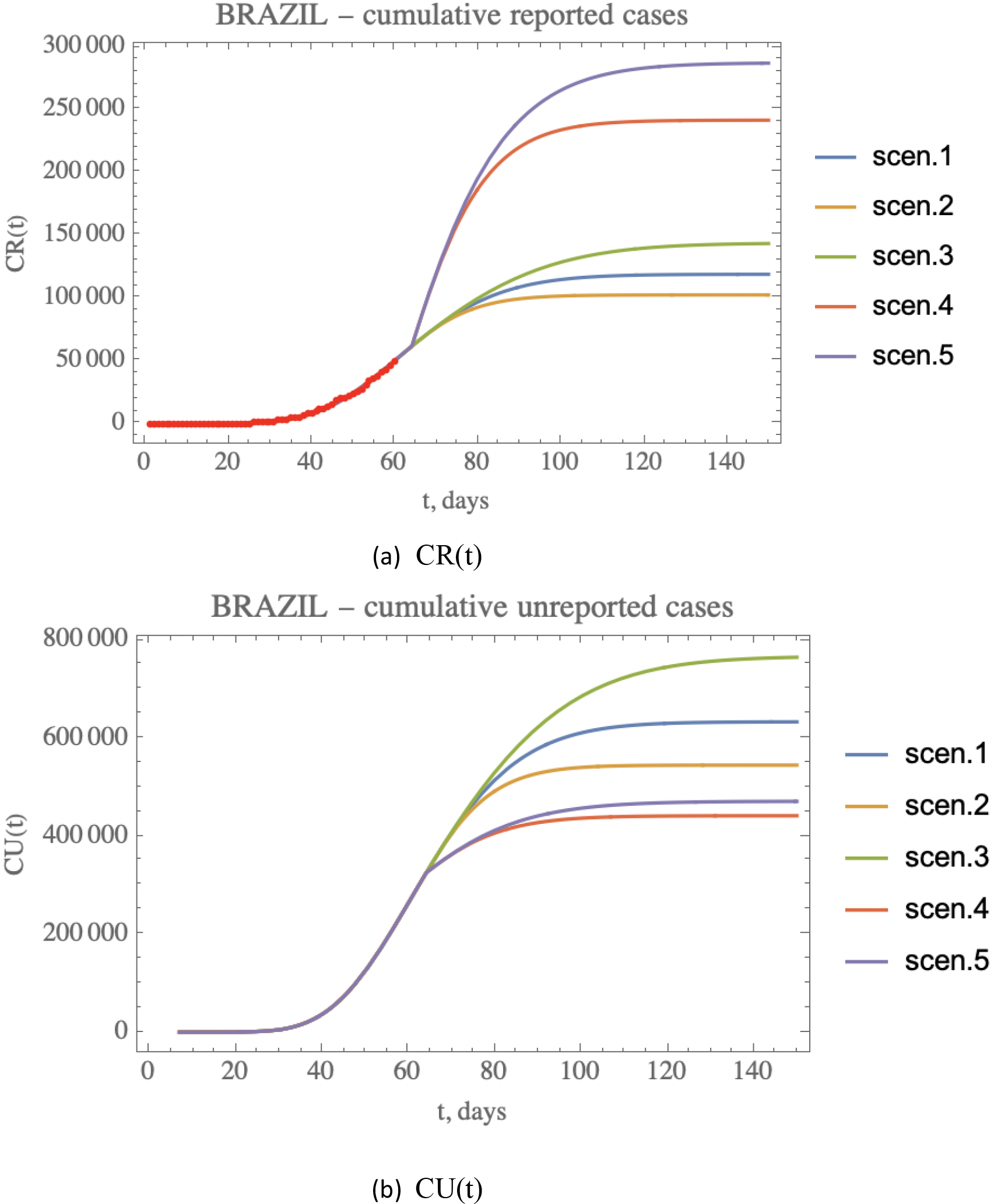
a,b –Comparative predictions for a) CR(t), and b) CU(t), for the five scenarios (i) to (v). (red dots in (a) show available data of CR(t) up to April 23^rd^).

## CONCLUSIONS

The present work implements a mixed analytical-statistical inverse problem analysis to the prediction of epidemics evolution, with focus on the COVID-19 progression in Brazil. A SIRU-type model is implemented for the direct problem solution, while a mixture of an analytical parametric estimation for the early phase epidemic exponential behavior with a Bayesian inference approach for a wider period, that encompasses the initial public health interventions to control the epidemics, are considered for the inverse problem analysis. The evolution of the COVID-19 epidemy in China is considered for validation purposes, by taking the first part of the dataset of accumulated reported infectious individuals to estimate parameters, and retaining the rest of the evolution data for direct comparison with the predicted results, with excellent agreement. Then, the same approach is applied to the Brazilian case, this time employing an initial portion of the available time series so far for the parametric estimates, and then offering a validation of the evolution prediction through the remaining dataset up to the date available at conclusion of this study (April 23rd). Also, some public health intervention measures are critically examined through five different scenarios, in addition to those already implemented, permitting the inspection of their impact on the overall dynamics of the disease proliferation. It was observed that a combination of social distancing and sanitary habits with a more intensive testing for isolation of symptomatic cases, could lead to the same overall control of the disease than a more strict social distancing intervention. Further improvement on the modelling is envisioned by enriching the model with latency effects, age structure discrimination, spatial demographic distribution dependence, and recovery factor differentiation among isolated and non-isolated patients.

## Data Availability

Data is available upon request from the corresponding author.

## ACKNOWLEDGEMENTS

The authors are deeply grateful to Dr. Tania Mattos Petraglia, MD, for the valuable information on COVID-19 pathology and treatment.

---

**Table A.1.** - Data for Brazil - accumulated reported cases, CR(t), and casualties.

| DATE | Day | Death | Infected | DATE | Day | Death | Infected |
| --- | --- | --- | --- | --- | --- | --- | --- |
| 24/02/2020 | 1 | 0 | 0 | 30/03/2020 | 36 | 163 | 4630 |
| 25/02/2020 | 2 | 0 | 1 | 31/03/2020 | 37 | 201 | 5717 |
| 26/02/2020 | 3 | 0 | 1 | 01/04/2020 | 38 | 242 | 6880 |
| 27/02/2020 | 4 | 0 | 1 | 02/04/2020 | 39 | 324 | 8044 |
| 28/02/2020 | 5 | 0 | 1 | 03/04/2020 | 40 | 363 | 9194 |
| 29/02/2020 | 6 | 0 | 2 | 04/04/2020 | 41 | 445 | 10360 |
| 01/03/2020 | 7 | 0 | 2 | 05/04/2020 | 42 | 486 | 11254 |
| 02/03/2020 | 8 | 0 | 2 | 06/04/2020 | 43 | 564 | 12183 |
| 03/03/2020 | 9 | 0 | 2 | 07/04/2020 | 44 | 686 | 14034 |
| 04/03/2020 | 10 | 0 | 3 | 08/04/2020 | 45 | 820 | 16188 |
| 05/03/2020 | 11 | 0 | 8 | 09/04/2020 | 46 | 954 | 18145 |
| 06/03/2020 | 12 | 0 | 13 | 10/04/2020 | 47 | 1068 | 19789 |
| 07/03/2020 | 13 | 0 | 19 | 11/04/2020 | 48 | 1140 | 20962 |
| 08/03/2020 | 14 | 0 | 25 | 12/04/2020 | 49 | 1223 | 22192 |
| 09/03/2020 | 15 | 0 | 25 | 13/04/2020 | 50 | 1328 | 23430 |
| 10/03/2020 | 16 | 0 | 34 | 14/04/2020 | 51 | 1532 | 25262 |
| 11/03/2020 | 17 | 0 | 52 | 15/04/2020 | 52 | 1757 | 28610 |
| 12/03/2020 | 18 | 0 | 77 | 16/04/2020 | 53 | 1947 | 30683 |
| 13/03/2020 | 19 | 0 | 151 | 17/04/2020 | 54 | 2141 | 33682 |
| 14/03/2020 | 20 | 0 | 151 | 18/04/2020 | 55 | 2347 | 36599 |
| 15/03/2020 | 21 | 0 | 200 | 19/04/2020 | 56 | 2462 | 38654 |
| 16/03/2020 | 22 | 0 | 234 | 20/04/2020 | 57 | 2575 | 40581 |
| 17/03/2020 | 23 | 1 | 346 | 21/04/2020 | 58 | 2741 | 43079 |
| 18/03/2020 | 24 | 4 | 529 | 22/04/2020 | 59 | 2906 | 45757 |
| 19/03/2020 | 25 | 7 | 640 | 23/04/2020 | 60 | 3313 | 49492 |
| 20/03/2020 | 26 | 11 | 970 |  |  |  |  |
| 21/03/2020 | 27 | 18 | 1178 |  |  |  |  |
| 22/03/2020 | 28 | 25 | 1546 |  |  |  |  |
| 23/03/2020 | 29 | 34 | 1924 |  |  |  |  |
| 24/03/2020 | 30 | 46 | 2247 |  |  |  |  |
| 25/03/2020 | 31 | 57 | 2433 |  |  |  |  |
| 26/03/2020 | 32 | 77 | 2985 |  |  |  |  |
| 27/03/2020 | 33 | 92 | 3417 |  |  |  |  |
| 28/03/2020 | 34 | 111 | 3904 |  |  |  |  |
| 29/03/2020 | 35 | 136 | 4256 |  |  |  |  |

